# Frequency & Relationship between Loneliness and Smartphone Addiction among Medical Students of Lahore

**DOI:** 10.1101/2024.02.16.24302853

**Authors:** Ammar Sohail

## Abstract

Smartphone addiction is a widespread and concerning issue wherein individuals excessively and compulsively use their smartphones. It’s characterized by habitual and prolonged phone usage, resulting in negative impacts on physical and mental health, strained personal relationships, increased fixation on the device, and a notable decrease in both personal productivity and overall quality of life. This study aims to find relationship between smartphone addiction and loneliness & prevalence of loneliness and smartphone addiction among medical students in Pakistan. This will also help to gauge the extent of related health issues and will lead to formulation of better guidelines for people suffering from smartphone addiction & loneliness thus improving the social and mental wellbeing of people from all age groups in Pakistani society.

## INTRODUCTION

Loneliness is a distressing experience that occurs when a person’s social relationships are perceived by that person to be less in quantity, and especially in quality, than desired. The experience of loneliness is highly subjective; an individual can be alone without feeling lonely and can feel lonely even when with other people (1). The world has become a global village, and people are more connected to each other than ever before. The problem that we are facing in current era is that this wave of technology has socially withdrawn people from each other. People are spending more times on their gadgets than actually interacting with each other in-person.

According to a global survey, about **33 percent** of adults experienced feelings of loneliness worldwide (2). The results shown in a study that **5**.**3 percent** of the participants (international students in China) were severely distressed by loneliness that may require professional help (3). The COVID-19 pandemic had a greater impact on the social and mental wellbeing of the people. The people are more prone to developing mental health issues due to psychological toll of COVID-19 pandemic on people’s lives (4). The pandemic also contributed to the loneliness among the masses due to movement restrictions, social distancing and lockdown measures. COVID-19 pandemic also contributed to increased screen time since most people were working from home and doing remote jobs. Mobile phone usage significantly increased among people of all age groups, especially medical students (5). A study conducted in twin cities of Pakistan suggested that the **60%** participants of study were suffering from smartphone addiction, out of which **57**.**3%** were males and **42**.**6%** were females. Smartphone addiction was found to be high among Pakistani adolescents (6). A study conducted at Bahauddin Zakariya University Multan (BZU) and Islamia University of Bahawalpur (IUB) showed that 28% university students are suffering from internet addiction, and 13.5% have high level of loneliness (7).

This study aims to find relationship between loneliness and smartphone addiction & prevalence of loneliness and smartphone addiction among medical students in Pakistan. These issues not only negatively impact learning of the students, but also jeopardize their physical and mental wellbeing. These issues are not being currently addressed in Pakistan at community level, and there are no policy guidelines for people suffering from loneliness and smartphone addiction. This study tends to improve the social and mental wellbeing of medical students in Pakistan by assessing the levels of loneliness and smartphone addiction & their interrelationship. This will also help to gauge the extent of related health issues and will lead to formulation of better guidelines for people suffering from smartphone addiction and loneliness thus improving both physical and mental well-being of people from all age groups in Pakistani society.

## Methodology

This study adopted an online survey as the main method of data collection. Cross sectional survey method used for this research. Data was collected through self-administered response forms via Google Forms.. Participants answered the questions anonymously. All analyses were done among these 414 participants.

### Sample size

Sample size was calculated to be 414, with 95% confidence interval. Medical colleges were selected via random sampling, while students were selected via systematic random sampling.

### Inclusion criteria

1. Medical students currently enrolled in MBBS & BDS programs for more than one year.
2. Students from both public and private medical colleges in Lahore are eligible to participate.

### Exclusion criteria

1. Students currently studying in first year.

### Statistical Analysis

Statistical analysis was performed by using Statistical Package for Social Sciences (SPSS), Version 27.0. Analyzed data by using descriptive statistics, in which Mean, Standard Deviation, and Chi-Square test for comparing mean of both University students from score of smartphone addiction and loneliness.

### Scales/Questionnaires

All participants were assessed using the UCLA Loneliness Scale, and the Smartphone Addiction Scale (SAS). Demographic characteristics were also assessed.

#### UCLA Loneliness Scale

The 10-item UCLA Loneliness Scale (e.g., ‘‘How often do you feel unhappy doing so many things alone?’’) on a four-point Likert scale (1 = never and 4 = always) was adopted to achieve a unidimensional measure of loneliness.26,67 The reliability Cronbach’s alpha was 0.87.

| Questions | Never | Rarely | Sometimes | Often |
| --- | --- | --- | --- | --- |
| How often do you feel lack of companionship? | 1 | 2 | 3 | 4 |
| How often do you feel that there is no one you can turn to? | 1 | 2 | 3 | 4 |
| How often do you feel you are no longer close to anyone? | 1 | 2 | 3 | 4 |
| How often do you feel your interests and ideas are not shared by those around you? | 1 | 2 | 3 | 4 |
| How often do you feel left out? | 1 | 2 | 3 | 4 |
| How often do you feel that your social relationships are superficial? | 1 | 2 | 3 | 4 |
| How often do you feel that no one really knows you well? | 1 | 2 | 3 | 4 |
| How often do you feel unhappy being so withdrawn? | 1 | 2 | 3 | 4 |
| How often do you feel people are around you but not with you? | 1 | 2 | 3 | 4 |

#### Smartphone addiction

To assess the intensity of smartphone addiction, a six-point Likert scale (1 = strongly disagree and 6 = strongly agree) was used on the 15-item SAS scale (e.g., ‘‘Missing planned work due to smartphone use’’).50 The reliability Cronbach’s alpha was 0.908.

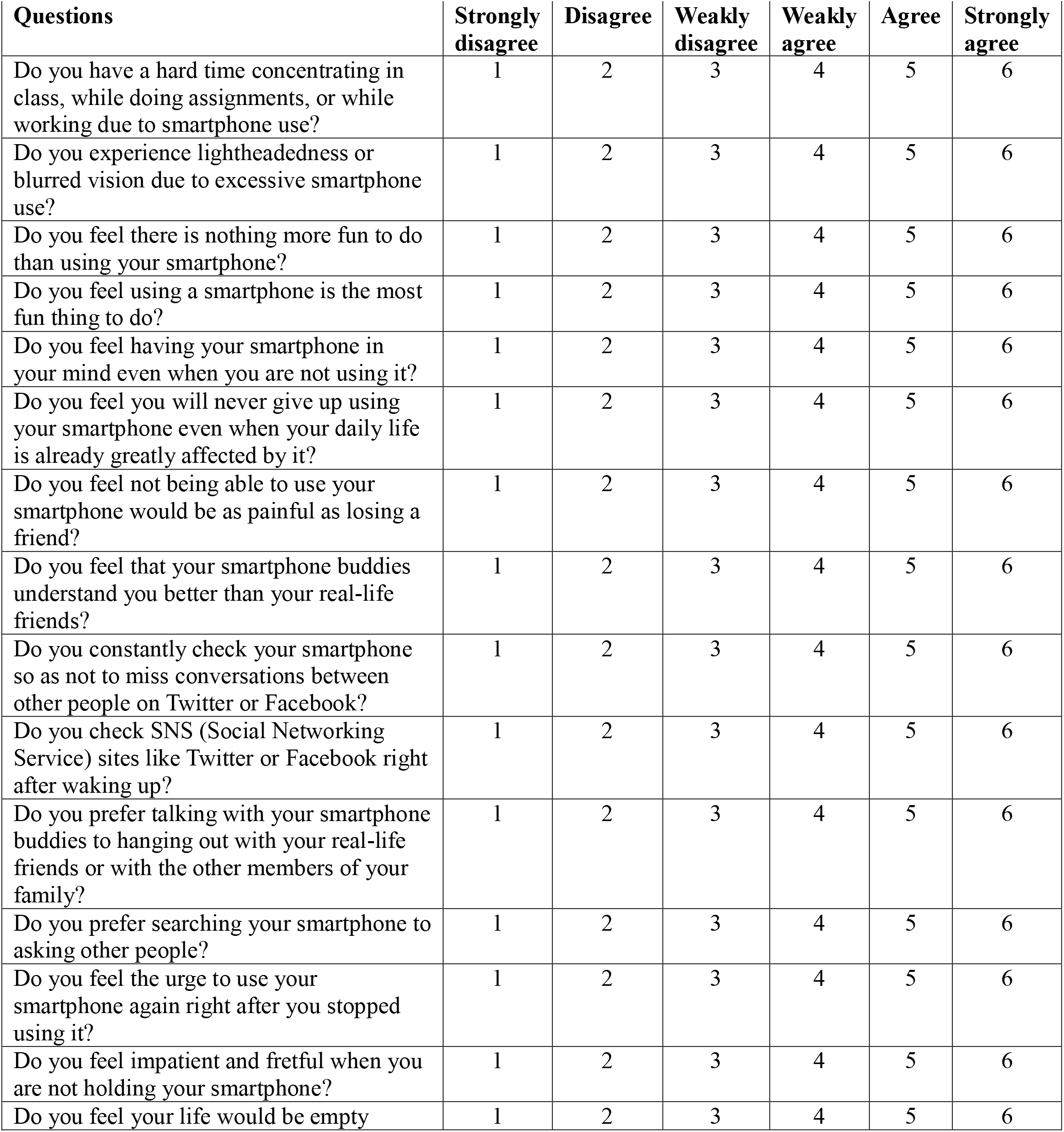
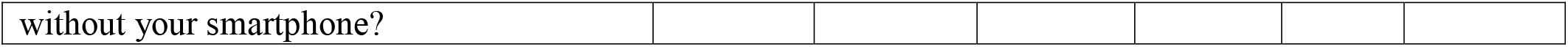

### Operational Definition of Variables

#### Smartphone Addiction

Smartphone addiction refers to a widespread and concerning issue wherein individuals excessively and compulsively use their smartphones. It’s characterized by habitual and prolonged phone usage, resulting in negative impacts on physical and mental health, strained personal relationships, increased fixation on the device, and a notable decrease in both personal productivity and overall quality of life.

#### Loneliness

The state of being alone in solitary isolation and isolates from the real world and deprives them of the sense of belonging and connection with real world contacts as feelings of emptiness without the companionship of others or alienated as a function of an inability to establish, maintain, and terminate relationships appropriately and interrupts real life relationships.

### Ethical Considerations

Permission for using the scale has been taken from the respective authors. A departmental permission has also been taken from where the data was collected through the participants. Informed consent taken from all respondents and debriefing the participants regarding research. All participants have filled questionnaires willingly; forced choice not applied on any respondents and not provides any reward or financial assistance to the participants.

## Results

Descriptive analyses were run for the total UCLA scores. The average score for loneliness for medical students is 26. Scores between 15 and 20 are considered a normal experience of loneliness, while scores above 30 indicate that a person is experiencing severe loneliness. The UCLA scores were calculated, with the total scores ranging from 10 to 40 (mean = 26.74, S.D = 7.099). As for levels of loneliness, 42 percent of the participants showed UCLA scores no more than 20 (little level of loneliness), 23.2 percent ranged between 21 and 30 (moderate level of loneliness), and 34.8 percent are above 30, experiencing severe loneliness.

Descriptive analyses were also run for the total scores of smartphone addiction. The total SAS scores were calculated, ranging from 15 to 80 (mean = 52.04, SD = 13.62). As for levels of smartphone addiction, scores below 20 is considered mild smartphone addiction, 23.2 percent of the participants showed SAS scores between 20 and 30 which showed moderate addiction, above scores 31 onwards i.e. 34.8 percent were in the severe smartphone addiction group. In addition, 30 percent of the participants who suffered from severe smartphone addiction were also experiencing severe loneliness, who might be in the need of professional help.

### Statistical analysis

To test the relationship among smartphone addiction and loneliness, Chi-Square test was run which showed significant relationship between smartphone addiction and loneliness among medical students in Lahore (p= <0.001).

Table 1 & 2 show that out of 414 medical students studying in different medical colleges 34.8 percent are suffering from severe loneliness whereas 42 percent of the population has little level of loneliness. Similarly, 29 percent have severe smartphone addiction whereas 20.8 percent have mild addiction.

**Table 1:** Frequency of Loneliness.

| Table 1: Frequency of Loneliness |  |  |
| --- | --- | --- |
|  | Frequency | Percent |
| Little | 174 | 42.02 |
| Moderate | 96 | 23.14 |
| Severe | 144 | 34.22 |
| Total | 414 | 100 |

**Table 2:** Frequency of Smartphone Addiction.

| Table 2: Frequency of Smartphone Addiction |  |  |
| --- | --- | --- |
|  | Frequency | Percent |
| mild | 86 | 20.77 |
| moderate | 208 | 50.28 |
| severe | 120 | 28.98 |
| Total | 414 | 100 |

The table 3 shows the descriptive statistics of all the variables concerned in the scales.

**Table 3:** Descriptive Statistics of all the Variables.

addiction whereas 20.8 percent have mild addiction. The table 3 shows the descriptive statistics of all the variables concerned in the scales.
| Table 3: Descriptive Statistics of all the Variables |  |  |
| --- | --- | --- |
|  | Loneliness | Smartphone Addiction |
| N | 414 | 414 |
| Mean | 26.744 | 52.0435 |
| Median | 27 | 54 |
| S.D. | 7.09924 | 13.62674 |

Out of 414 participants, 210 (50.7%) are males and 204 (49.3%) are females. The Table 4 shows the gender wise distribution of smartphone addiction among medical students (p = 0.301), whereas Table 5 represents the gender wise distribution of Loneliness (p = 0.013);

**Table 4:**
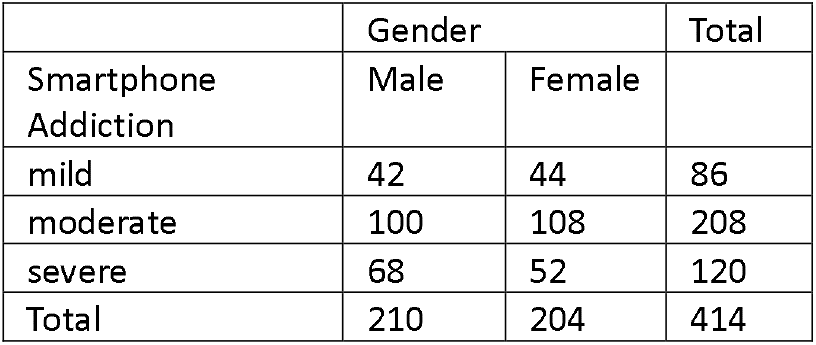

| Table 4: | Gender |  | Total |
| --- | --- | --- | --- |
| Smartphone Addiction | Male | Female |  |
| mild | 42 | 44 | 86 |
| moderate | 100 | 108 | 208 |
| severe | 68 | 52 | 120 |
| Total | 210 | 204 | 414 |

**Table 5:**

Out of 414 participants, 210
| Table 5: |  | Gender |  | Total |
| --- | --- | --- | --- | --- |
|  |  | Male | Female |  |
| Loneliness | mild | 88 | 86 | 174 |
|  | moderate | 60 | 36 | 96 |
|  | severe | 62 | 82 | 144 |
| Total |  | 210 | 204 | 414 |

Result of Table 5 shows that level of loneliness is greater in male as compared to female hence we conclude that males feel more loneliness as compared to females.

The Table 7 shows the comparison of trends of Loneliness among different colleges of Lahore:

**Table 7:**

| Table 7: |  | Loneliness |  |  | Total |
| --- | --- | --- | --- | --- | --- |
|  |  | mild | moderate | severe |  |
| Name of Medical College | SKZMDC | 34 | 18 | 34 | 86 |
|  | FJMU | 32 | 12 | 32 | 76 |
|  | AIMC | 12 | 6 | 6 | 24 |
|  | KEMU | 11 | 8 | 4 | 23 |
|  | LMDC | 28 | 10 | 12 | 50 |
|  | AMC | 12 | 1 | 14 | 27 |
|  | CMH | 6 | 2 | 0 | 8 |
|  | AVMDC | 10 | 13 | 8 | 31 |
|  | SMDC | 14 | 10 | 7 | 31 |
|  | SHMDC | 15 | 16 | 27 | 58 |
| Total |  | 174 | 96 | 144 | 414 |

The table 7 shows significant findings of different levels of loneliness students are suffering from (p = 0.03), however it is worth mentioning that different medical colleges have different response ratio of questionnaire, so actual results may vary.

Table 8 shows the trends of smartphone addiction among medical colleges of Lahore:

**Table 8:**

| Table 8: |  | Smartphone Addiction |  |  | Total |
| --- | --- | --- | --- | --- | --- |
|  |  | mild | moderate | severe |  |
| Name of Medical College | SKZMDC | 22 | 44 | 20 | 86 |
|  | FJMU | 28 | 40 | 8 | 76 |
|  | AIMC | 0 | 16 | 8 | 24 |
|  | KEMU | 7 | 6 | 10 | 23 |
|  | LMDC | 12 | 26 | 12 | 50 |
|  | AMC | 1 | 15 | 11 | 27 |
|  | CMH | 0 | 8 | 0 | 8 |
|  | AVMDC | 4 | 12 | 15 | 31 |
|  | SMDC | 1 | 14 | 16 | 31 |
|  | SHMDC | 11 | 27 | 20 | 58 |
| Total |  | 86 | 208 | 120 | 414 |

The table 8 shows significant findings of different levels of smartphone addiction students are suffering from (p = <0.01), however it is worth mentioning that different medical colleges have different response ratio of questionnaire, so actual results might be different. Due to lack of responses from certain medical colleges, significant results are unable to be displayed here.

According to the difference of accommodation, either day scholar or Hostellite, there are different levels of smartphone addiction and loneliness among these students. Out of 414 students, 230 (55.6%) are Hostellites and 184 (44.4%) are day scholars. Chi-Square tests show there is little to no significant relationship between accommodation & loneliness (p = 0.233) and accommodation & smartphone addiction (p = 0.899) represented by Tables 9 & 10 respectively.

**Table 9:**

| Table 9: |  | Loneliness |  |  | Total |
| --- | --- | --- | --- | --- | --- |
|  |  | mild | moderate | severe |  |
| Accommodation | Hostellite | 90 | 60 | 80 | 230 |
|  | Day Scholar | 84 | 36 | 64 | 184 |
| Total |  | 174 | 96 | 144 | 414 |

**Table 10:**

| Table 10: |  | Smartphone Addiction |  |  | Total |
| --- | --- | --- | --- | --- | --- |
|  |  | mild | moderate | severe |  |
| Accommodation | Hostellite | 46 | 116 | 68 | 230 |
|  | Day Scholar | 40 | 92 | 52 | 184 |
| Total |  | 86 | 208 | 120 | 414 |

**Table 11:**
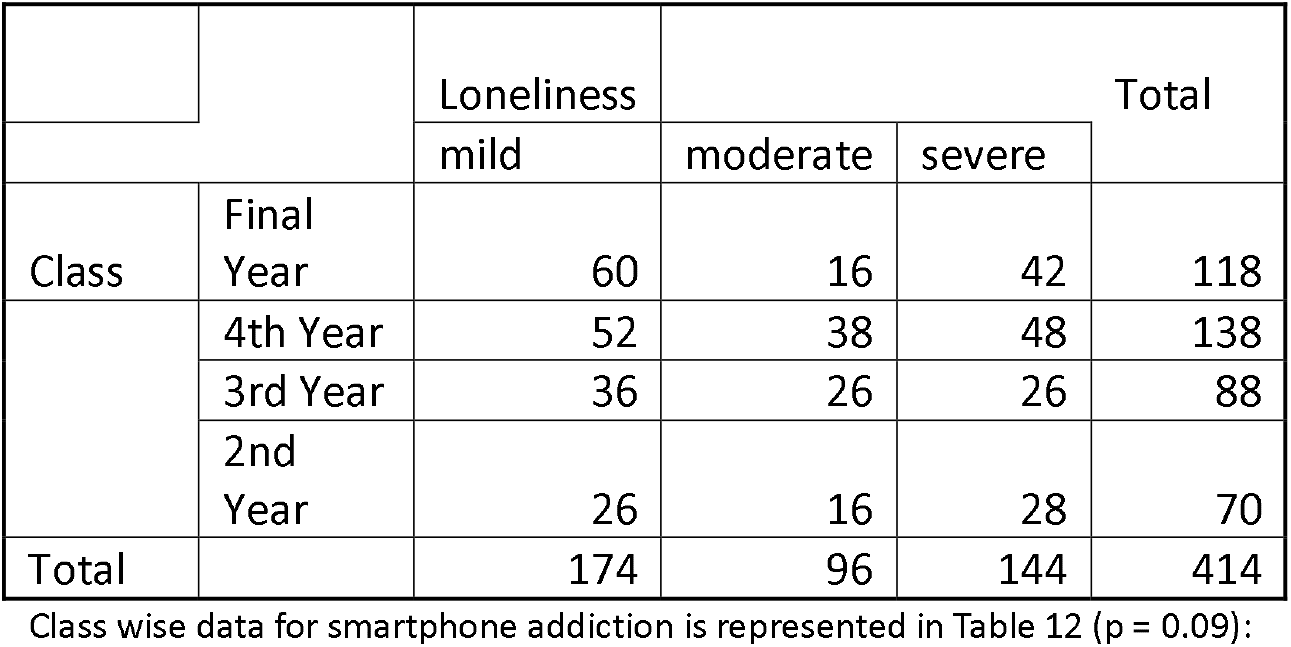

**Table 12:**

Class wise data for smartphone addiction is represented in Table 12 ( $p = 0.09$ ):
| Table 12: |  | SFC8 |  |  | Total |
| --- | --- | --- | --- | --- | --- |
|  |  | mild | moderate | severe |  |
| Class | Final Year | 22 | 66 | 30 | 118 |
|  | 4th Year | 22 | 78 | 38 | 138 |
|  | 3rd Year | 24 | 34 | 30 | 88 |
|  | 2nd Year | 18 | 30 | 22 | 70 |
| Total |  | 86 | 208 | 120 | 414 |

## Discussion

This research is critical as both loneliness and smartphone addiction are rising concerns that significantly affect individuals’ mental and physical health, personal relationships, productivity, and overall quality of life. The emergence of technology and the COVID-19 pandemic have further exacerbated these issues, making them particularly relevant for the youth and students, who are increasingly relying on smartphones for social interaction, education, and entertainment. The research aimed to assess the relationship and prevalence of loneliness and smartphone addiction among medical students in Lahore, to understand the extent of related health issues, and to establish guidelines to alleviate these problems. The study employed an online survey using Google Forms, where 414 medical students from various colleges in Lahore participated. The UCLA Loneliness Scale and the Smartphone Addiction Scale (SAS) were utilized to measure loneliness and smartphone addiction, respectively. A significant portion of the participants displayed high levels of loneliness and smartphone addiction. Out of 414 students, 34.8% experienced severe loneliness, and 28.98% had severe smartphone addiction. There was a significant relationship between loneliness and smartphone addiction, indicating that higher smartphone addiction is associated with increased feelings of loneliness. The sample size was 414, with students from both public and private medical colleges. Students were predominantly in their second year and above, with a balanced distribution of males and females. Data analysis revealed that loneliness and smartphone addiction are prevalent among medical students, with severe loneliness found in 34.8% of participants, while 28.98% suffer from severe smartphone addiction. The relationship between smartphone addiction and loneliness was statistically significant. The study concludes that smartphone addiction and loneliness are significantly interrelated among medical students in Lahore, with a considerable percentage experiencing severe problems in both areas. This highlights the need for professional help and the development of effective strategies to address these issues among students to improve their mental and social well-being.

## Conclusion

This study concluded that 120 (28.98%) have severe smartphone addiction out of 414 medical students of Lahore. Smartphone addiction has significant relationship with loneliness. Students suffering from smartphone addiction also feel more loneliness as compare to those who are not. These students might need professional help to make good use of their capabilities and potentials.

## Data Availability

All data produced in the present work are contained in the manuscript. Manuscript contains all the data of research.

